# Global burden of maternal hypertensive disorder from 1990 to 2021

**DOI:** 10.1101/2025.04.01.25325061

**Authors:** Bin Liu, Xiying Huang, Ziyong Hao, Jian Wang, Yiting Fan, Qin Shao, Ruogu Li, Ben He, Lisheng Jiang

## Abstract

**Background:** Maternal hypertensive disorder (MHD) is a significant global health concern, affecting 5% to 10% of pregnant women. We aimed to systematically estimate the global, regional, and national burden and temporal trends of MHD from 1990 to 2021.

**Methods:** Data including the incidence, deaths, and disability-adjusted life years (DALYs), along with their age-standardized rates (ASRs) of MHD were obtained from the Global Burden of Disease Study 2021 and stratified by age and the sociodemographic index (SDI). An age-period cohort (APC) model was employed to clarify the impacts of age, period, and cohort. Decomposition and frontier analyses were used to assess the influencing factors and disease disparities, respectively.

**Results:** In 2021, there were 18,050,085 (95% uncertainty interval [UI]: 15,356,124 to 21,519,204) incident cases, 38,147 (95% UI: 31,879 to 46,096) deaths and 2,469,637 (95% UI: 2,083,398 to 2,958,213) DALYs cases globally. The ASRs of incidence, deaths, and DALYs significantly decreased from 1990 to 2021. The burden of MHD was negatively correlated with SDI. Age-specific analysis revealed the highest burden in the 20-34 age group, with increasing trends in the 35-54 age group in high SDI regions. The APC model highlighted significant period and cohort effects, with improvements in high-middle SDI regions.

**Conclusions:** Although the global burden of MHD has decreased, significant disparities persist, particularly in low SDI regions, requiring targeted interventions such as strengthening healthcare infrastructure and international cooperation to address the burden. In high SDI regions, managing lifestyle risk factors is also crucial in pregnant women with advanced age.

## Introduction

Maternal hypertensive disorders (MHD) is a collective term encompassing various hypertensive conditions linked with pregnancy, including gestational hypertension, preeclampsia, eclampsia, and chronic hypertension exacerbated by preeclampsia.^1^ It is the second leading cause of maternal mortality after obstetric hemorrhage, affecting approximately 5% to 10% of pregnant women globally.^2, 3^ Moreover, 20% to 30% of women with a history of hypertensive disorders in pregnancy are likely to experience recurrence in subsequent pregnancies.^4, 5^ Compared with non-MHD pregnant women, those with MHD have a 2.4-fold increase in developing hypertension within 10 years postpartum, and elevate the long-term risks of cardio-cerebrovascular diseases.^6^

Although the global burden of MHD has improved over the past 30 years, significant disparities persist across regions and age groups. In Africa and the Caribbean, MHD mortality and Disability-adjusted life years (DALYs) rates remain high, with projected substantial increases in deaths and DALYs among adolescent pregnant women over the next 25 years.^7^ Additionally, the incidence of MHD varies globally and is closely associated with socioeconomic status.^8^ This indicates that the epidemiological characteristics of MHD are complex, with lower socioeconomic regions facing greater challenges in prevention and control, necessitating targeted interventions.

Previous studies have reported the global burden of MHD, however, the timeliness of data is crucial for public health decision-making and resource allocation.^7^ Therefore, it is necessary to update these data. In this study, we aim to accurately assess the global and regional trends in the burden of MHD based on the latest Global Burden of Disease (GBD) 2021 estimates. We have expanded the scope of previous research to report the incidence, mortality, and DALYs of MHD in 204 countries and regions from 1990 to 2021 and further analyzed these data by age, sociodemographic index (SDI), and age-period-cohort (APC) models.

## Methods

### Data acquisition and download

The GBD 2021 report utilizes the most recent epidemiological data and refined standardization methodologies to comprehensively assess the global burden of 371 diseases, injuries, and 88 risk factors across 204 countries and 21 territories stratified by age and sex. In this study, the incidence, mortality, and DALYs for MHD, along with their 95% uncertainty interval (UI), are derived from the GBD 2021 data.^9^ Additionally, the study incorporates the SDI, an indicator that quantifies the level of sociodemographic progress of a country or region based on income, education, and fertility metrics. ^10^ In GBD 2021, MHD is categorized based on the international classification of diseases, the 9th edition (ICD-9), which includes codes 642-642.9, and the 10th edition (ICD-10), which includes codes O10-O16.9.^11^

## Statistical analysis

### Descriptive analysis

Using the GBD 2021 estimates, a thorough evaluation was performed to measure the burden of MHD. This assessment included the analysis of incidence, mortality, and DALYs, as well as age-standardized rate (ASR) at global, regional, and national levels. Furthermore, the study explored demographic factors affecting the burden of MHD, analyzing the spread of the disease’s impact across various age groups. We calculated the age-standardized incidence rate (ASIR), age-standardized mortality rate (ASMR), and age-standardized DALYs rate (ASDR) based on the GBD World Standard Population using the following formula: 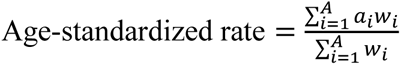 according to previous report, where a_i_ is the age-specific rate and w_i_ is the weight in the same age subgroup of the chosen reference standard population (in which i denotes the i^th^ age class) and A is the upper age limit.^12^

### Trend analysis

To measure the trend changes in the ASR over a specified time interval, our study calculated the estimated annual percentage change (EAPC). A linear model was constructed by fitting a regression line to the natural logarithm of ASR, represented as y=α+β+ε, in which y=ln (ASR) and x = calendar year. The EAPC is calculated using the formula EAPC=100 × (*e*^*β*^−1). The model also facilitates the calculation of the 95% confidence interval (CI). ^13^ When the estimated EAPC and the lower limit of its 95% CI are both positive, the ASR is indicative of an increasing trend. Conversely, when the estimated EAPC and the upper limit of its 95% CI are both negative, the ASR declines. If the 95% CI overlaps, the ASR is regarded as stable.

### Decomposition analysis and frontier analysis

This study employs the decomposition method invented by Das Gupta to quantify the specific impacts of aging, population growth, and epidemiological changes on the global burden of MHD. ^14, 15^ To thoroughly assess the relationship between the burden of MHD and the level of sociodemographic development, we utilized data from 1990 to 2021 to construct a frontier analysis model based on the ASIR, ASMR, ASDR and SDI. This model aims to more accurately reveal the potential improvement space in the ASR of MHD that a country or region might achieve. ^16^

### APC model

The study utilized an age-period-cohort analysis to assess the individual influences of age, period, and cohort factors on the incidence rate, deaths rate, and DALYs rate of MHD in different regions. In this study, we concentrated on estimating the net drift, the local drift, the longitudinal age curve, and the period rate ratio (PRR) and cohort rate ratio (CRR). Net drift represents the overall logarithmic tendency across different periods and birth cohorts. Local drift refers to the average annual percentage change (AAPC) for each age group after considering period and cohort effects. The longitudinal age curve is the age-specific longitudinal rates adjusted for cohort and period effects. The period (cohort) rate ratio represents the relative risk of a period (or cohort) compared to a reference period (or cohort), adjusted for age and non-linear cohort (or period) effects. In the APC analysis, we divided the data into sequential 5-year spans ranging from 1990 to 2021 and into consecutive 5-year age brackets from 10-14 up to 50-54. Due to the difficulty of segmenting the 1990 and 1991 data into complete 5-year intervals, these two years were excluded from the analysis to ensure consistency and accuracy in the study.

In the present study, Pearson correlation was also used to assess the relationships between SDI and ASIR, ASMR, and ASDR. All statistical analyses were conducted using R (version 4.3.3).

## Result

### MHD Incidence and Its Change Trend

In 2021, a significant decrease was observed in the global incidence of MHD compared with 1990. The incidence cases increased from 15,662,895 (95% UI, 12,935,146–19,254,514) to 18,050,085 (95% UI, 15,356,124–21,519,204), however, the ASIR decreased from 554.4 per 100,000 population (95% UI, 461.4–675.4) to 461.9 per 100,000 population (95% UI, 392.7–551.7), with an EAPC of −0.51 (95% CI, −0.56–-0.45). The low SDI regions had the most substantial MHD burden with the incidence cases increasing from 4,139,304 (95% UI, 3,487,342–4,867,143) in 1990 to 6,749,882 (95% UI, 5,727,623–7,955,523) in 2021 (**Table 1**). From 1990 to 2021, the ASIR demonstrated an annual downward trend across all SDI regions except the high-middle SDI regions. In the low-middle SDI regions, the EAPC was −1.83 (95% CI: −1.99, −1.68), showing the most significant decreasing trend (**Table 1** and **Figure 1A**). At the regional level, the ASIR decreased in all regions from 1990 to 2021, except for Eastern Europe and Western Europe. Eastern Europe observed the most significant increase, with an EAPC of 1.12 (95% CI, 0.65–1.59) (**Table 1**). In 2021, India, Nigeria, and Pakistan had the highest incidence cases, while Tokelau, Niue, and San Marino had the fewest (**Table S3** and **Figure 2**). South Sudan, Niger, and Chad exhibited the highest ASIR, whereas the Republic of Korea, Canada, and Luxembourg shown the lowest in 2021 (**Table S3** and **Figure S1**). From 1990 to 2021, Kazakhstan had the highest EAPC for ASIR (**Table S3** and **Figure S2)**.

**Figure 1.**
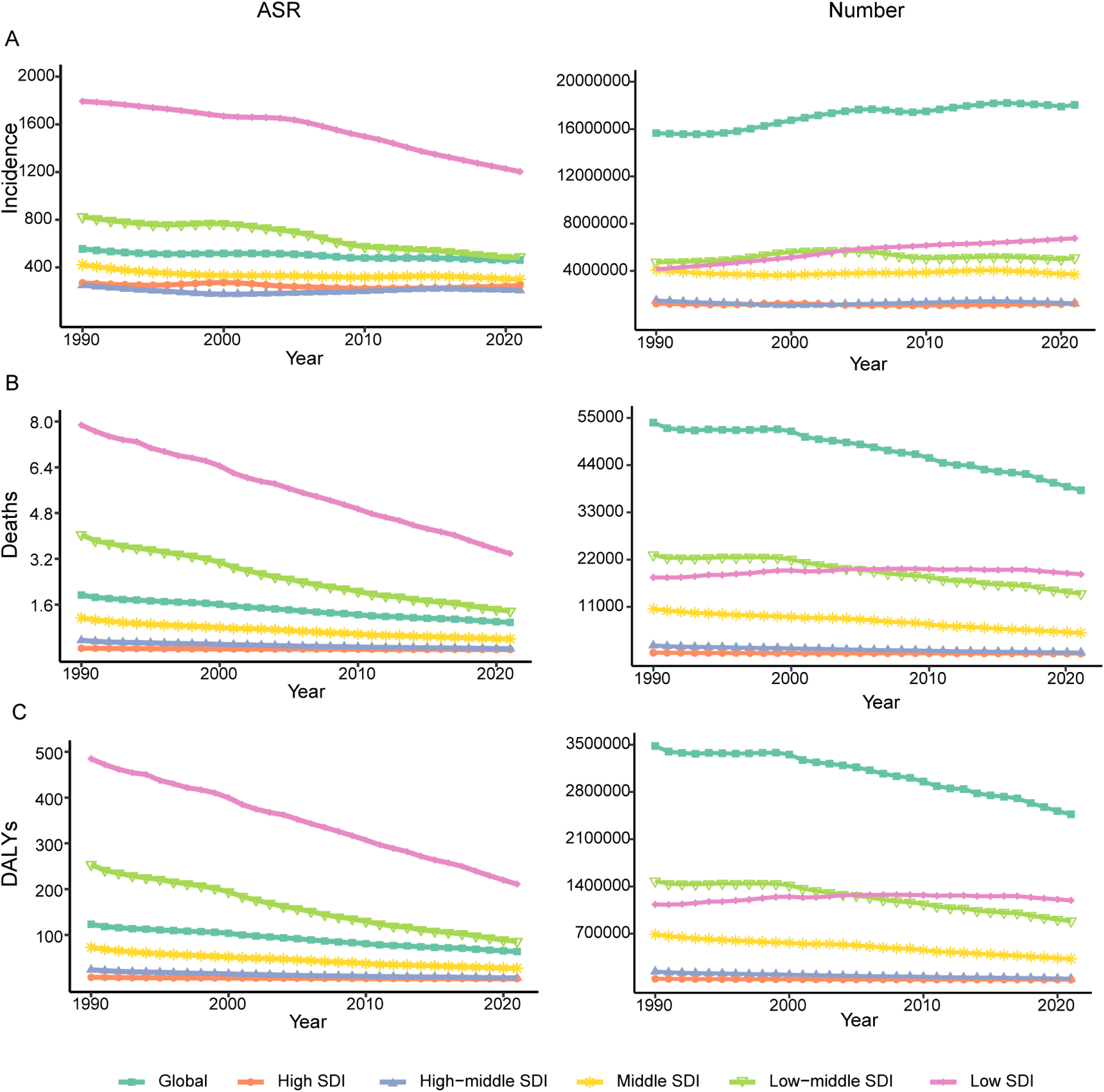
The trend of MHD burden across different SDI regions from 1990 to 2021. (A) ASIR and number of incidence; (B) ASMR and number of deaths; (C) ASDR and number of DALYs. MHD: maternal hypertensive disorder; SDI: sociodemographic index; ASR: age-standardized rate; ASIR: age-standardized incidence rate; ASMR: age-standardized mortality rate; ASDR: age-standardized DALYs rate; DALYs: disability-adjusted life years.

**Figure 2.**
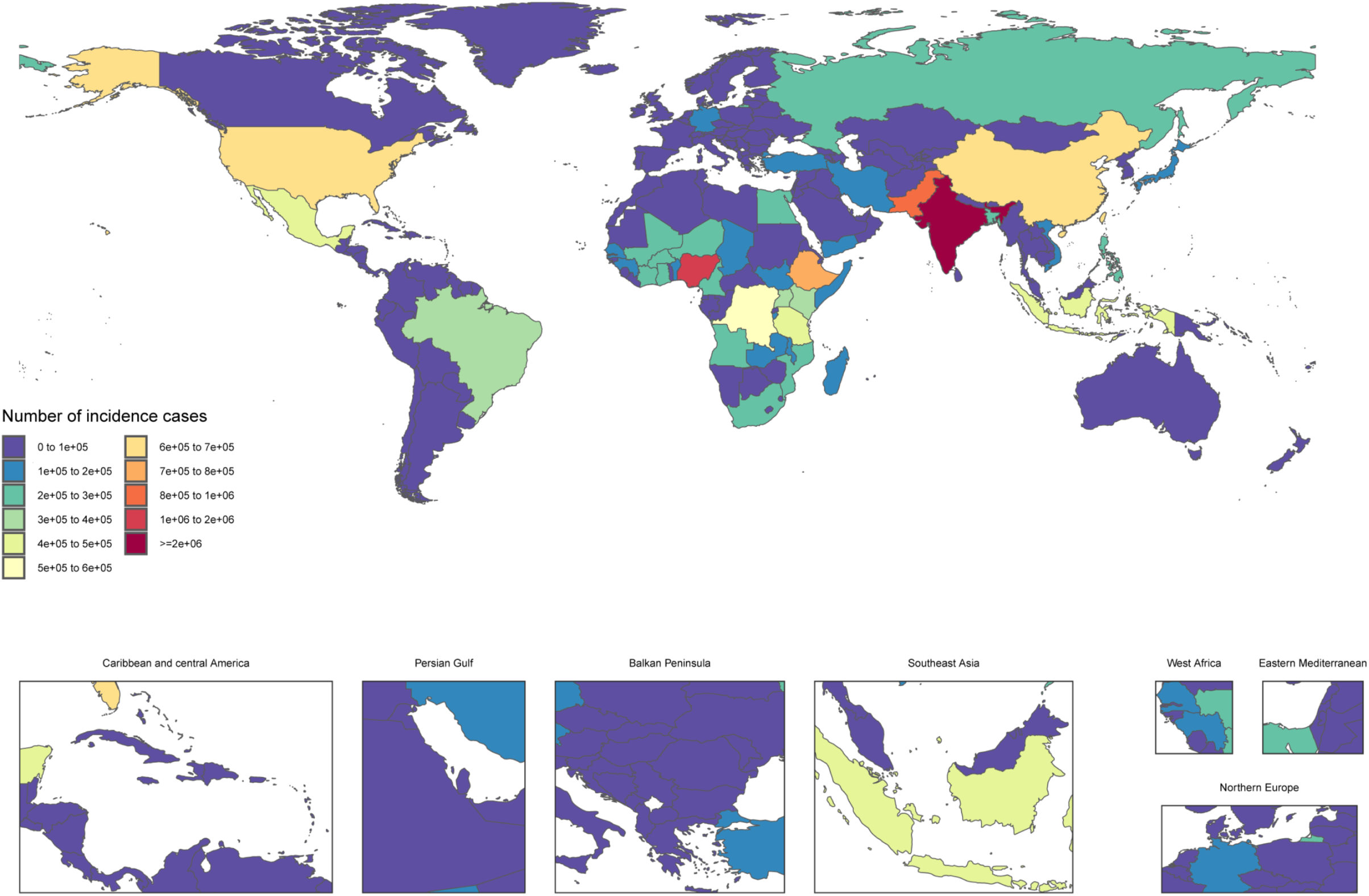
Number of incidence cases of MHD in 2021 by country and territory. MHD: maternal hypertensive disorder.

**Table 1.**
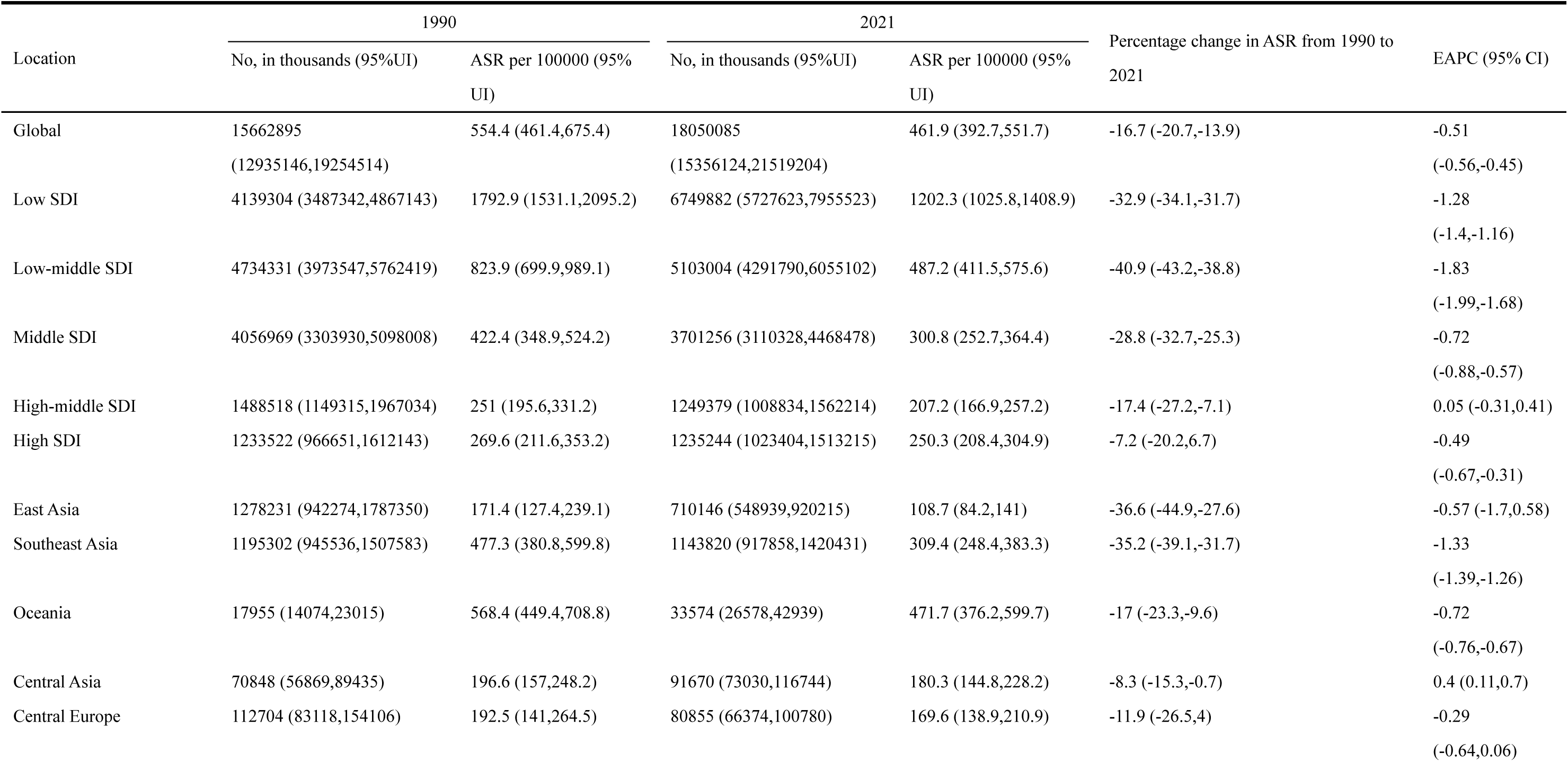
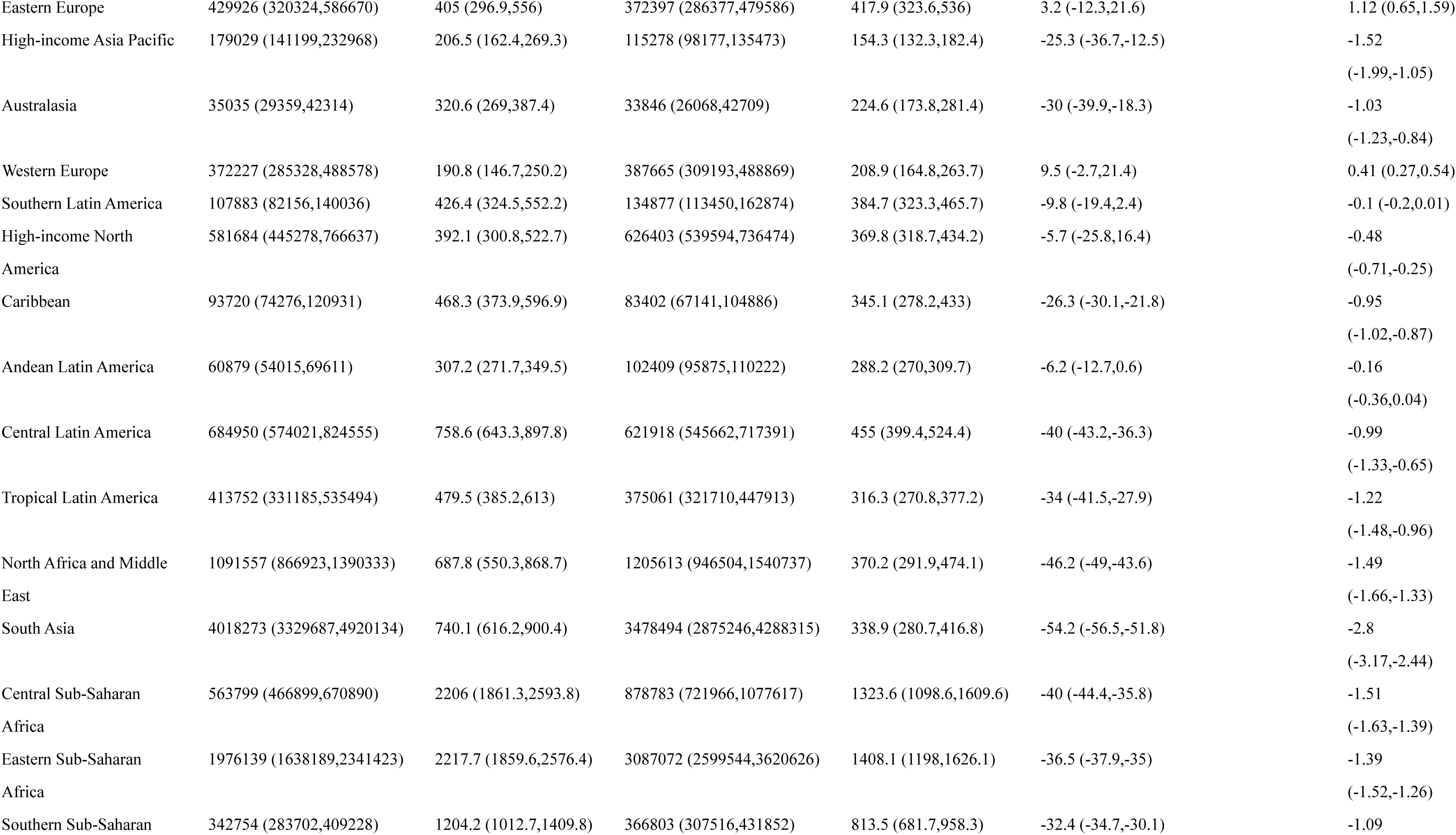
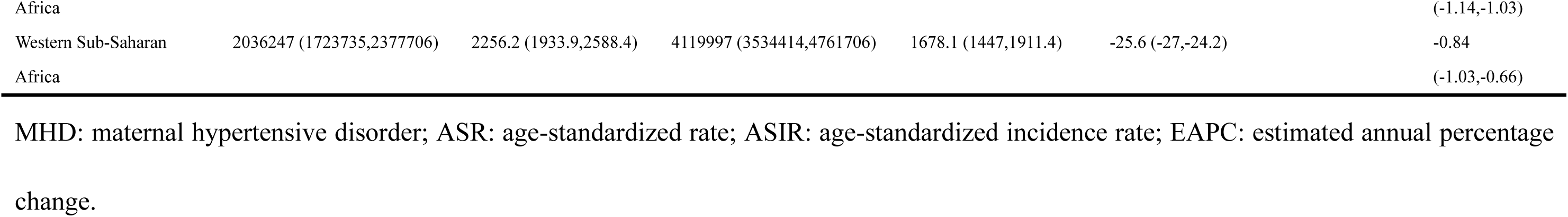
The incidence cases and ASIR of MHD, alongside the percentage change in ASR per 100,000 population and the EAPC by location from 1990 to 2021.

### MHD Deaths and Its Change Trend

There was a significant decrease in mortality, dropping from 53,894 (95% UI, 47,676–59,819) in 1990 to 38,147 (95% UI, 31,879–46,096) in 2021(**Table S1**). The ASMR also decreased from 1.9 per 100,000 population (95% UI, 1.7–2.2) in 1990 to 1.0 (95% UI, 0.8–1.2) in 2021(**Table S1**). Only low SDI regions had a mortality rise (1990: 17,872 [95% UI, 15,191-20,320]; 2021: 18,589 [95% UI, 15,234-22,700]) (**Table S1** and **Figure 1B**). All 5 SDI regions saw an ASMR decline, but the lowest was in low SDI regions, with an EAPC of - 2.6 (95% CI, - 2.7 to - 2.5) (**Table S1** and **Figure 1B**). At regional level, South Asia had the most deaths (1990: 20,401 [95% UI, 17,206–23,202]; 2021: 11,967 [95% UI, 8,863–15,766]), but its ASMR shown a significant decrease (EAPC: - 3.92 [95% CI, - 4.11 to - 3.74]) (**Table S1**). The Caribbean had the largest ASMR increase, with an EAPC of 0.8 (95% CI, 0.58 to 1.02) (**Table S1**). At national level, India, Nigeria, and Pakistan had the highest mortality in 2021 (5,630, 5,527, and 5,464, respectively) (**Table S1** and **Figure S3**). South Sudan and Chad showed the highest ASMR in 2021(**Table S4** and **Figure S4**). From 1990 to 2021, Guam had the highest EAPC for ASIR (**Table S4** and **Figure S5**).

### MHD DALYs and Its Change Trend

The DALYs of MHD had a significant decrease, from 3,479,885 (95% UI, 3,085,883–3,873,201) in 1990 to 2,469,637 (95% UI, 2,083,398-2,958,213) in 2021, and the ASDR also witnessed a significant decline, with an EAPC of −2.1 (95% CI, −2.15 to −2.04) (**Table S2**). Only low SDI regions’ DALYs increased, from 1,134,178 (95% UI, 960,803–1,289,414) in 1990 to 1,194,635 (95% UI, 983,739–1,454,751) in 2021(**Table S2** and **Figure 1B**). All 5 regions’ ASDR is decreasing, with the high SDI regions seeing the smallest drop, with an EAPC of −1.7 (95% CI, −1.81 to −1.6) (**Table S2** and **Figure 1B**). At regional level, South Asia had the most DALYs in 1990 (1,315,573 [95% UI, 1,116,161–1,497,912]) and 2021 (746,671 [95% UI, 559,140–976,528]). Only the Caribbean saw a rise in ASDR over this time (EAPC: 0.75, [95% CI, 0.53-0.96]) (**Table S2**). At national level, India, Indonesia, and Bangladesh had the most DALYs cases in 1990, with 736,898, 300,869, and 292,455, respectively. By 2021, the top countries were Nigeria, India, and Pakistan, with DALYs of 356825, 353143, and 336427, respectively (**Table S5** and **Figure S6**). Chad and South Sudan showed the highest ASDR in 2021 (**Table S5** and **Figure S7**). From 1990 to 2021, Zimbabwe had the highest EAPC for ASIR (**Table S5** and **Figure S8**).

### Correlations between SDI and the burden of MHD

From 1990 to 2021, the ASIR, ASMR, and ASDR of MHD at the regional level were inversely related to SDI level (R =-0.75, P < 0.001; R =-0.81, P < 0.001; R =-0.81, P < 0.001, respectively) (**Figure 3**, **Figure S9** and **Figure S10**). In 2021, the ASIR, ASMR, and ASDR of MHD at the national level were also negatively associated with SDI (R =-0.78, P < 0.001; R =-0.77, P < 0.001; R =-0.78, P < 0.001, respectively) (**Figure 4**, **Figure S11**, and **Figure S12**). South Sudan had the highest global ASIR and ASMR [2337.9 (95% UI: 2015.4 to 2688.9); 6.9 (95% UI: 4.1 to 11.3) per 100 000 population], while the Republic of Korea had the lowest ASIR and ASDR [35 (95% UI: 28.2 to 42.4); 0.9 (95% UI: 0.6 to 1.2) per 100 000 population] (**Table S3**-**S5**, **Figure 4**, **Figure S11** and **Figure S12**). The highest ASDR [447.1 (95% UI: 297.4 to 639.9) per 100 000 population] was in Chad, and Slovenia had the lowest ASMR [0 (95% UI: 0 to 0) per 100 000 population] (**Table S4**, **Table S5**, **Figure S11** and **Figure S12**). Between 1990 and 2021, Libya shown the largest decline in ASIR [-68.1% (95% UI: −72.4 to −63.3)], while Jordan saw the most significant decrease in ASMR and ASDR [-96.8% (95% UI: −98.3 to −94) and −95.7% (95% UI: −97.5 to −92.2)] (**Table S3**-**S5**). Ecuador had the largest increase in ASIR [51.4% (95% UI: 48.1 to 54.5)], and Guam witnessed the most substantial rise in ASMR and ASDR [42.5% (95% UI: −24.5 to 152) and 20.6% (95% UI: −13.6 to 72.6)] (**Table S3**-**S5**).

**Figure 3.**
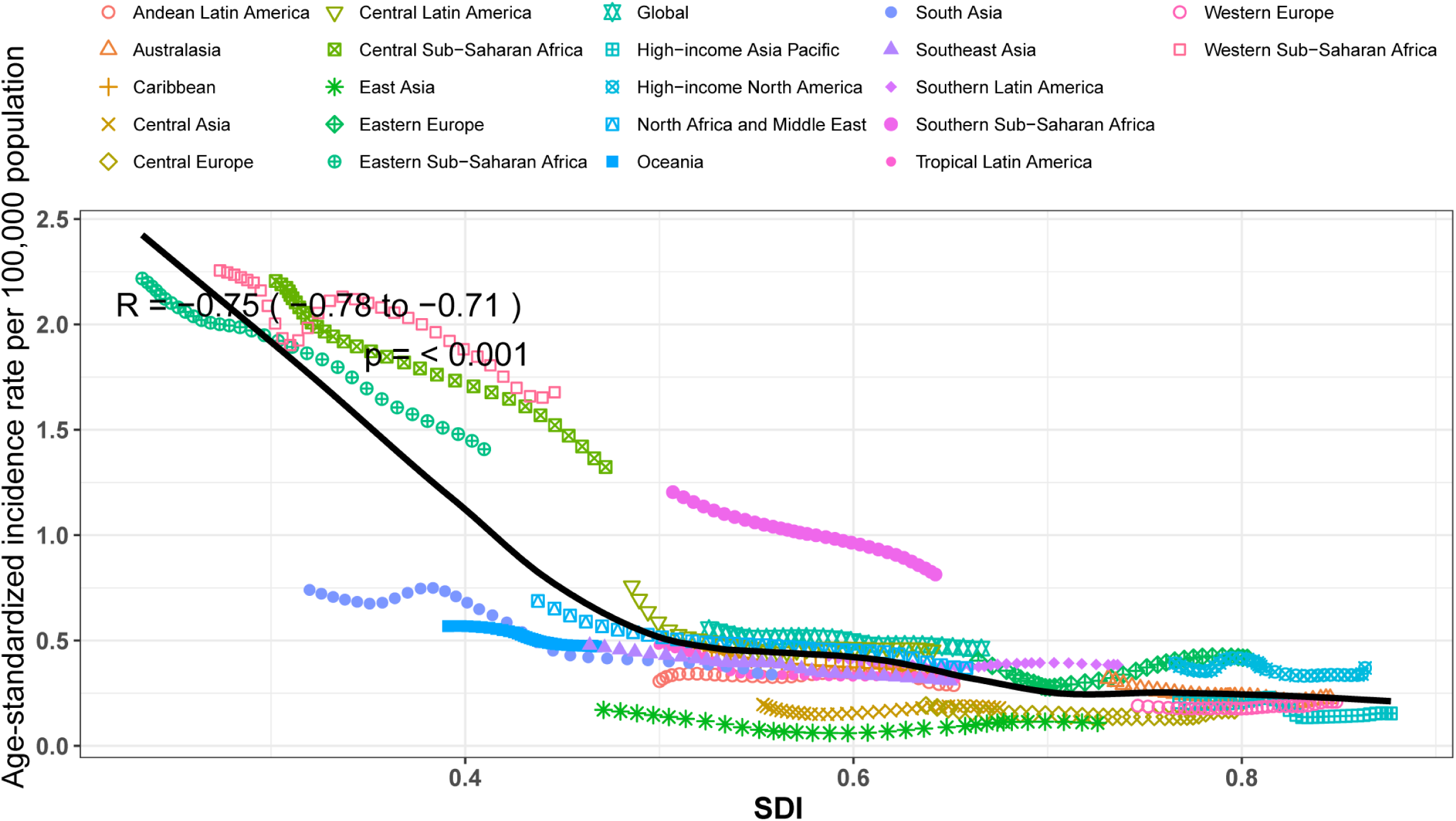
Age-standardized incidence rate for MHD for Global Burden of Disease regions by SDI from 1990–2021. MHD: maternal hypertensive disorder; SDI: sociodemographic index.

**Figure 4.**
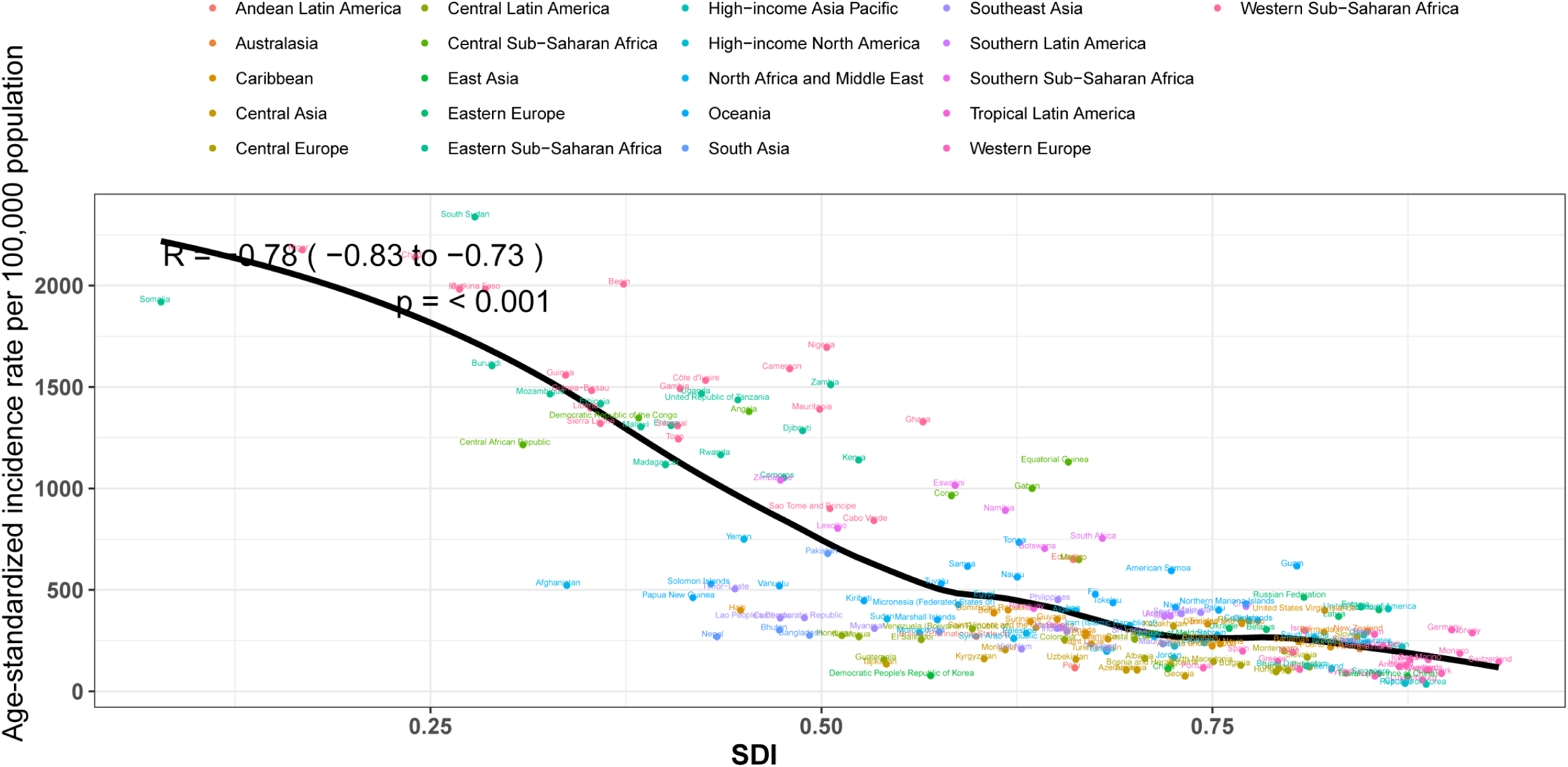
Age-standardized incidence rate of MHD for 204 countries and territories by SDI in 2021. MHD: maternal hypertensive disorder; SDI: sociodemographic index.

### Age-specific patterns of MHD in global and 5 SDI regions

Our study revealed changes in the burden of MHD among different age groups in different regions from 1990 to 2021. The ASIR, ASMR, and ASDR are generally decreasing in 5 SDI regions except for the 35-54 age group (**Figure 5** and **Table S6**-**S8**). The lowest disease burden was observed in the 10-19 age group, while the highest was in the 20-34 age group (**Figure 5**, **Figure S13**, and **Table S6**-**S8**). In high SDI and high-middle SDI regions, however, the ASIR in the 35-54 age group has increased, with EAPC of 2.34 (95% CI 2.25-2.43) and 2.52 (95% CI 2.04-3.01), respectively (**Figure 5A**, **Figure S14A**, and **Table S6**). Only in high SDI regions, the ASDR in the 35-54 age group showed a slight upward trend, with an EAPC of 0.12 (95% CI 0.02-0.22) (**Figure 5C**, **Figure S14C**, and **Table S8**).

**Figure 5.**
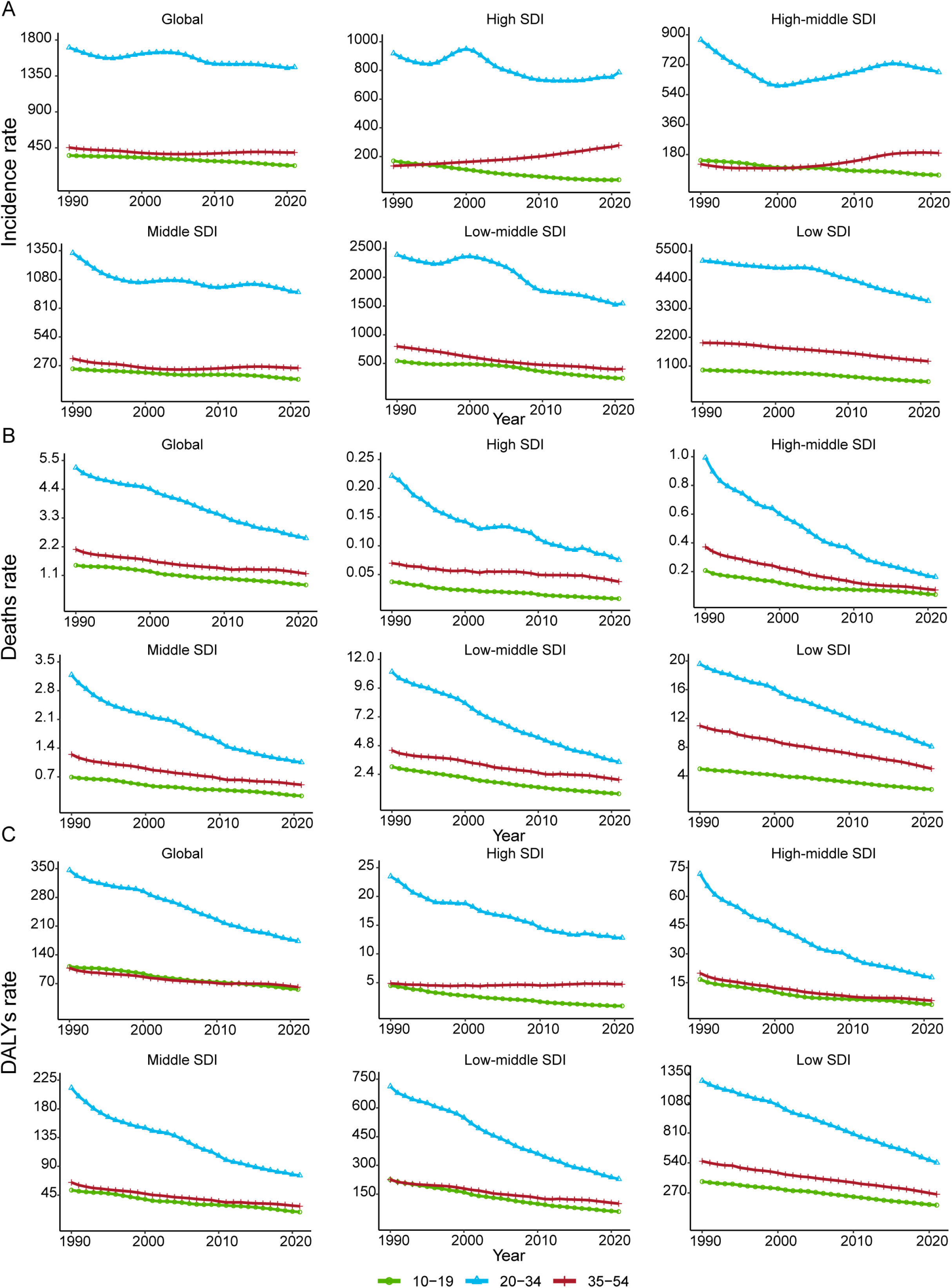
The rate of MHD burden across different age groups in different regions from 1990 to 2021. (A) Incidence rate; (B) Death rate; (C) DALYs rate. MHD: maternal hypertensive disorder; SDI: sociodemographic index; DALYs: disability-adjusted life years.

### Trends in APC for ASIR, ASMR, and ASDR in MHD

In terms of ASIR, high SDI and high-middle SDI regions show an upward trend around age 35. High-middle SDI regions have a net drift over 0 (EAPC=0.05, 95% CI: −0.31 to 0.41), while low-middle SDI regions have the highest decline (EAPC=-1.83, 95% CI: −1.99 to −1.68) (**Figure S15A**). In the 30 - 40 age group, high SDI regions have a higher ASIR than high-middle and middle SDI regions (**Figure S15B**). Low-middle SDI regions have the greatest drop in PRR and CRR, whereas high-middle and high SDI regions perform poorly, with high-middle SDI regions showing a slight PRR and CRR increase (**Figure S15C**-**S15D**).

The APC model for ASMR is similar to the ASDR, so we will not go into details here (**Figure S16**-**S17**). Regarding DALYs, the net drift for global and 5 SDI regions is below zero, indicating a decline in ASDR after adjusting for period and cohort effects from 1992 to 2021 (**Figure S17A**). The most significant decrease is observed in the high-middle SDI regions (EAPC=-4.19, 95% CI: −4.27 to −4.10), while the high SDI regions show the least pronounced decline, with an upward trend in ASDR observed in the 35-54 age group (**Figure S17A**). The ASDR increases until about 25 years old and then declines, with the following hierarchy within the same age group: low SDI > low-middle SDI > global average > middle SDI > high-middle SDI > high SDI (**Figure S17B**). From 1992 to 2021, the PRR generally decreased, with the largest decline in the high-middle SDI regions and the smallest in the high SDI regions (**Figure S17C**). Compared to the cohort from 1967 to 1977, the CRR was higher before 1967 and lower after 1977, with the high-middle SDI regions also showing a significant decrease and the high SDI regions exhibiting the smallest decline throughout this trend (**Figure S17D**).

### Decomposition Analysis

This study uses decomposition analysis to divide the shifts in the burden of MHD into three main elements: epidemiological changes, population growth, and population aging (**Figure S18** and **Table S9**). From 1990 to 2021, epidemiological changes were the dominant factor in the significant decrease in global MHD mortality (204.93%) and DALYs (199.32%) (**Figure S18B**-**S18C** and **Table S9**). Conversely, population growth was the main contributor to the significant rise in global MHD incidence (291.24%) (**Figure S18A** and **Table S9**). In all SDI regions, the changes in mortality and DALYs were generally consistent with the global changes, except for the low SDI regions, where the changes were opposite to the global trends due to the impact of population growth (**Figure S18B-S18C** and **Table S9**). Epidemiologic changes had a positive impact on improving the global burden of MHD in all SDI regions (**Figure S18B-S18C** and **Table S9**). In addition, the impact of population growth on the rising incidence of MHD was greatest in low SDI and low-middle SDI regions (**Figure S18A** and **Table S9**).

### Frontier analysis

The frontier analysis reveals distinct trends in MHD’s ASRs as SDI values change from 1990 to 2021 across 204 countries and territories (**Figure S19A**-**S21A**). For incidence, ASIR declines with SDI rising from 0.0 to 1.0, shown by a density shift from light to dark shades over the years (**Figure S19A**). Similarly, ASMR and ASDR decrease with increased SDI level, indicating that the socio-economic developing reduces the burden of MHD (**Figure S20A**-**S21A**). In 2021, the visual representations delineated clear distinctions among countries and territories (**Figure S19B**-**S21B**). For ASIR, 15 countries, including South Sudan and Chad, had much higher rates, placing them far from the frontier (**Figure S19B**, **Table S10**). For ASMR and ASDR, South Sudan and Chad were farther from the frontier (**Figure S20B**-**S21B**, **Table S11**-**S12**).

## Discussion

This study, based on the GBD 2021 data, systematically evaluated the trends in the incidence, mortality, and DALYs of MHD from 1990 to 2021. The findings revealed that: First, the ASIR, ASMR, and ASDR of MHD showed a significant decline globally, but the burden of MHD remained high in the low SDI regions, likely due to population growth, despite the largest decline in ASIR observed in these regions. Second, the ASIR, ASMR, and ASDR of MHD were significantly negatively correlated with the SDI, indicating that higher levels of social development contribute to reducing the burden of MHD. Third, although the disease burden was lowest in the 10-19 age group and highest in the 20-34 age group across all SDI regions, the ASIR for the 35-54 age group showed an increasing trend in high SDI and high-middle SDI regions, and a slight increase in ASDR was also observed in the same age group in high SDI regions. These findings underscore the importance of strengthening targeted public health interventions globally, particularly in regions with lower SDI, to reduce the burden of MHD and improve maternal health effectively.

Our findings reveal that although the global MHD cases increased from 15.66 million in 1990 to 18.05 million in 2021, the ASIR significantly declined (from 554.4 to 461.9 per 100,000 population). This paradoxical phenomenon suggests that population growth may have masked the actual effectiveness of disease prevention and control efforts. Notably, while the case number in low and low-middle SDI regions increased over the 32 years, these regions also exhibited the most pronounced annual decline in ASIR (EAPC: −1.28 and −1.83, respectively), indicating potential progress in prevention and control measures. However, population growth partially offsets these achievements, which is supported by our decomposition analysis. Previous studies have reported an increase in the global burden of maternal diseases (particularly MHD) after 2020.^17^ Consistent with these findings, our study also observed a rise in both case numbers and ASIR of MHD in low-middle and high SDI regions after 2020, which may be associated with the increased pressure on healthcare systems during the pandemic of Corona Virus Disease 2019 (COVID-19). Additionally, existing research highlights that ongoing conflicts and resulting population displacement in regions such as Eastern Europe have had profound impacts on maternal and child health.^18^ Simultaneously, the high prevalence of metabolic disorders, including hypertension and diabetes, in Eastern Europe may have further contributed to this trend.^19^ Although Western Europe boasts advanced healthcare systems, factors such as increased immigration, economic austerity measures, and cuts in healthcare funding may have strained medical services.^20^ These factors collectively contributed to the observed increase in ASIR in these regions during the study period.

In this study, we observed that the trends in deaths and DALYs for MHD were highly consistent, showing a significant decline from 1990 to 2021. Notably, this downward trend persisted even during the COVID-19 pandemic, which may be attributed to the positive impact of advancements in medical technology and increased attention from healthcare professionals toward MHD.^21^ However, the number of deaths and DALYs in low SDI regions continued to rise, highlighting the severe challenges faced by resource-limited areas, where population growth may have further exacerbated the burden. Additionally, the disease burden in certain regions remains concerning. Specifically, we found that the ASR of mortality and DALYs due to MHD in the Caribbean exhibited a persistent upward trend, consistent with previous studies.^7^ At the same time, the burden in high-income North America showed no significant improvement. Numerous studies have indicated that the uneven distribution of healthcare resources, exposure to chlordecone, and the higher risk of MHD among individuals of Caribbean descent compared to their white counterparts may be key factors contributing to the increased burden of MHD in the Caribbean region.^22–24^ Additionally, the high prevalence of iron-deficiency anemia in high-income North America may also significantly exacerbate this phenomenon.^25^ These regions urgently require proactive intervention to address this pressing public health challenge.

Our study demonstrates a significant negative correlation between the ASIR, ASMR, and ASDR of MHD and the SDI level at regional and national levels. This finding indicates that although the global burden of MHD has improved compared to the past, low SDI regions continue to bear a disproportionately high burden, which aligns with previous research.^7, 26, 27^ Specifically, the incidence, mortality, and DALYs of MHD in Africa are significantly higher than in other regions. Previous statistical data and research analyses have revealed that approximately 22.1% of maternal deaths in sub-Saharan Africa are attributable to MHD.^2, 28^ In Central, Eastern, and Western Africa, MHD is the second most common cause of maternal mortality after obstetric hemorrhage.^29, 30^ However, this phenomenon is closely linked to the insufficient quality of maternal healthcare resources, as most African countries exhibit low coverage of early prenatal diagnosis for MHD, posing a significant challenge to improving adverse maternal outcomes caused by MHD.^31–33^ Additionally, African women frequently face risk factors like early or multiple pregnancies, advanced age, anemia, infections, and socioeconomic issues, which collectively drive up the region’s MHD burden.^34^ While high-middle SDI countries have advanced medical resources, rapid economic, industrial, and urban changes, along with globalization, have triggered major shifts in lifestyle and diet.^35^ These shifts in metabolic risk factors may have contributed to the mild increase in the EAPC of ASIR in these regions. For countries with higher SDI, focusing on preconception care and education to promote healthy lifestyles may help reduce the incidence of MHD.

Furthermore, our frontier analysis reveals the existence of significant disparities in the burden of MHD globally. Data from 2021 indicate that the burden of MHD in higher-SDI regions is substantially lower than in lower-SDI regions. Even though effective antihypertensive drugs are widely available and global guidelines emphasize strict blood pressure control, hypertension management is still inadequate, especially in low- and middle-income countries (LMICs).^36, 37^ While access to antihypertensive drugs provides a critical means to alleviate the global burden of MHD, barriers such as limited healthcare access, low awareness of hypertension, poor treatment adherence, and socioeconomic challenges hinder effective blood pressure control.^38^ A previous study revealed that among 44 LMICs, 74% of individuals with hypertension had their blood pressure measured, 39% were diagnosed, 30% received treatment, and only 10% achieved blood pressure control.^39^ In contrast, high SDI countries demonstrate superior hypertension control, suggesting that their strategies could serve as effective models globally.^37^ These strategies include community-based interventions, enhancing public awareness of hypertension, improving treatment adherence, and widespread implementation of updated guidelines.^40^ Higher levels of socioeconomic development are often associated with better healthcare infrastructure, improved access to medical services, and higher health literacy among populations. Therefore, promoting economic development and optimizing healthcare resource allocation in low SDI countries are critical to reduce these disparities and improve public health outcomes. To achieve this, strengthening international cooperation to facilitate the sharing of best practices, providing technical assistance, and mobilizing healthcare infrastructure resources are essential.

Our study further investigates the distribution of disease burden among pregnant women across different age groups. We analyzed data across five SDI levels based on the three categories of pregnant populations defined by the World Health Organization (WHO). Consistent with previous studies,^7^ women aged 20-34 years, who represent the primary childbearing age group, were identified as the major contributors to the burden of MHD. This finding underscores the importance of enhancing prenatal screening and health education for women of childbearing age. In contrast, the burden of adolescent pregnancies is the lowest, likely due to recent interventions and policy support, such as enhanced sexual education and increased access to contraceptive services.^41^ Furthermore, our study revealed a significant negative correlation between disease burden across all age groups and SDI levels, suggesting that higher income, improved education, better nutritional status, and enhanced access to healthcare resources associated with increased SDI may collectively mitigate the burden of MHD. Notably, in high SDI regions, the ASIR and ASDR for women aged 35-54 years exhibited an upward trend, which may be closely linked to prevalent lifestyle risk factors in high SDI countries, such as higher obesity rates, sedentary behavior, high-sodium diets, psychological stress, increased prevalence of advanced maternal age, and delayed childbearing due to the widespread use of assisted reproductive technologies. This paradoxical phenomenon highlights the need for high SDI regions to strengthen health management for older pregnant women to address their unique health challenges.

This study systematically examines the age, period, and cohort effects on the burden of MHD. Consistent with previous research,^27^ we observed a consistent age effect across different SDI regions, where the risk of MHD initially increased and then decreased with age, with women aged 20-29 identified as the highest-risk group. Additionally, these risks showed a significant negative correlation with SDI levels, suggesting that high SDI regions may effectively reduce the burden of MHD due to better healthcare resources and socioeconomic conditions. This finding indicates that women of childbearing age, at the peak of reproductive activity, face an elevated risk of pregnancy-related complications, while developed regions may mitigate this risk through more robust healthcare systems and social support. Interestingly, the risk associated with advanced maternal age exhibited a declining trend, which contrasts with the findings of most studies.^42, 43^ Although this observation contradicts previous research, some studies have reported that while advanced maternal age is a risk factor for gestational diabetes mellitus and cesarean delivery, it does not significantly increase the risk of MHD.^44, 45^

In terms of deaths and DALYs, the period effects indicate that the relative risk changes across all SDI regions globally followed a similar trend, with significant declines observed. This phenomenon may be attributed to the continuous improvements in healthcare and public health systems in recent years. Notably, the smallest decline occurred in high SDI regions, which is likely due to the already low disease burden in high SDI regions, making further substantial reductions challenging. Cohort effects also revealed similar relative risk changes across all SDI regions, with significant improvements, particularly in high-middle SDI regions, which exhibited the largest declines. This suggests that high-middle SDI regions have successfully reduced MHD-related mortality and improved adverse outcomes in recent years, offering valuable lessons for other regions. However, in terms of incidence of MHD from the APC analysis, unlike the significant declines in other SDI regions, high SDI and high-middle SDI regions made little progress in reducing the incidence. This phenomenon may be closely related to the widespread use of advanced MHD screening methods and unhealthy lifestyle factors in developed regions.

However, this study has several limitations. First, the reliance on the GBD 2021 database may be constrained by underreporting or diagnostic standard variations in some regions. Second, while SDI reflects macro-level socioeconomic status, it cannot capture micro-level factors such as healthcare accessibility or cultural practices. Third, the analysis was conducted at the SDI regional level without exploring country-specific differences, limiting the applicability of the findings to national contexts. Additionally, the GBD 2021 study did not provide subtype data for MHD, preventing an analysis of the global burden by MHD type.

## Conclusion

In summary, our study reveals that although the global burden of MHD has declined from 1990 to 2021, significant inequalities persist at regional, national and age levels, highlighting the need for more targeted prevention and control strategies. High SDI regions exhibit the lowest burden of MHD, while low and low-middle SDI regions bear the heaviest burden. Therefore, promoting economic development and optimizing healthcare resources in lower SDI regions is crucial, whereas higher SDI regions must remain vigilant against emerging risks associated with modern lifestyles. Additionally, health management and interventions targeting women of childbearing age should be prioritized in MHD prevention efforts.

## Perspectives

Our study reveals significant progress in reducing the burden of MHD and highlights persistent disparities and emerging challenges. The findings imply that higher SDI countries can focus on addressing lifestyle-related risk factors and optimizing screening strategies for older pregnant women, lower SDI countries urgently need enhanced economic development, improved healthcare resource allocation, and expanded access to prenatal care. Future research should examine the factors driving the rise in MHD incidence in higher SDI regions among women aged 35-54 and explore targeted interventions. Additionally, it should investigate the long-term effects of metabolic changes due to globalization in high-middle SDI countries. We need to adapt public health strategies to different sociodemographic contexts and foster international cooperation to share best practices and mobilize resources for reducing the global burden of MHD.

## Data Availability

Publicly available datasets were analyzed in this study. This data can be obtained from the IHME website at https://vizhub.healthdata.org/gbd-results/.

## Acknowledgments

We sincerely thank the contributors of the GBD 2021 and the Institute for Health Metrics and Evaluation (IHME) for their invaluable work. The study funders did not participate in the study’s design, data collection, analysis, interpretation, or report writing. L. Jiang and B. He designed the study. B. Liu and X. Huang analyzed the data, performed statistical analyses, and drafted the initial manuscript. L. Jiang and B. He checked and corrected the statistical analyses. Z. Hao, J. Wang, Y. Fan, Q. Shao, and R. Li modified the initial manuscript. All authors reviewed the drafted manuscript for critical content and approved the final version of the manuscript. The corresponding authors (L. Jiang and B. He) attest that all listed authors meet authorship criteria and that no others meeting the criteria have been omitted.

## Non-standard Abbreviations and Acronyms

MHD: maternal hypertensive disorder
DALYs: disability-adjusted life years
ASRs: age-standardized rates
SDI: sociodemographic index
APC: age-period cohort
UI: uncertainty interval
EAPC: estimated annual percentage change
GBD: Global Burden of Disease
ICD: international classification of diseases
ASR: age-standardized rate
ASIR: age-standardized incidence rate
ASMR: age-standardized mortality rate
ASDR: age-standardized DALYs rate
CI: confidence interval
PRR: period rate ratio
CRR: cohort rate ratio
AAPC: average annual percentage change
COVID-19: Corona Virus Disease 2019
LMICs: low- and middle-income countries
WHO: World Health Organization

## Supplemental Material

**Table S1.** The deaths cases and ASMR of MHD, alongside the percentage change in ASR per 100,000 population and the EAPC by location from 1990 to 2021.

**Table S2.** The DALY cases and ASDR of MHD, alongside the percentage change in ASR per 100,000 population and the EAPC by location from 1990 to 2021.

**Table S3.** The incidence cases and ASIR of MHD in 1990 and 2021, along with the percentage change in the ASR per 100,000 population and the EAPC by location.

**Table S4.** The deaths cases and ASMR of MHD in 1990 and 2021, along with the percentage change in the ASR per 100,000 population and the EAPC by location.

**Table S5.** The DALY cases and ASDR of MHD in 1990 and 2021, along with the percentage change in the ASR per 100,000 population and the EAPC by location.

**Table S6.** Age-specific of the incidence cases and ASIR of MHD in 1990 and 2021, along with the percentage change in the ASR per 100,000 population and the EAPC by location.

**Table S7.** Age-specific of the deaths cases and ASMR of MHD in 1990 and 2021, along with the percentage change in the ASR per 100,000 population and the EAPC by location.

**Table S8.** Age-specific of the DALYs cases and ASDR of MHD in 1990 and 2021, along with the percentage change in the ASR per 100,000 population and the EAPC by location.

**Table S9.** Decomposition analysis for MHD burden by different regions from 1990 to 2021.

**Table S10.** Frontier of MHD incidence and effective difference by location in 2021.

**Table S11.** Frontier of MHD deaths and effective difference by location in 2021.

**Table S12.** Frontier of MHD DALYs and effective difference by location in 2021.

**Figure S1.** Age-standardized incidence rate of MHD in 2021 by country and territory. MHD: maternal hypertensive disorder.

**Figure S2.** EAPC in incidence of MHD by country and territory from 1990 to 2021. MHD: maternal hypertensive disorder; EAPC: estimated annual percentage change.

**Figure S3.** Number of deaths cases of MHD in 2021 by country and territory. MHD: maternal hypertensive disorder.

**Figure S4.** Age-standardized deaths rate of MHD in 2021 by country and territory. MHD: maternal hypertensive disorder.

**Figure S5.** EAPC in deaths of MHD by country and territory from 1990 to 2021. MHD: maternal hypertensive disorder; EAPC: estimated annual percentage change.

**Figure S6.** Number of DALYs cases of MHD in 2021 by country and territory. MHD: maternal hypertensive disorder; DALYs: disability-adjusted life years.

**Figure S7.** Age-standardized DALYs rate of MHD in 2021 by country and territory. MHD: maternal hypertensive disorder; DALYS: disability-adjusted life years.

**Figure S8.** EAPC in DALYs of MHD by country and territory from 1990 to 2021. MHD: maternal hypertensive disorder; EAPC: estimated annual percentage change; DALYs: disability-adjusted life years.

**Figure S9.** Age-standardized deaths rate for MHD for Global Burden of Disease regions by SDI from 1990–2021. MHD: maternal hypertensive disorder; SDI: sociodemographic index.

**Figure S10.** Age-standardized DALYs rate for MHD for Global Burden of Disease regions by SDI from 1990–2021. DALYs: disability-adjusted life years; MHD: maternal hypertensive disorder; SDI: sociodemographic index.

**Figure S11.** Age-standardized deaths rate of MHD for 204 countries and territories by SDI in 2021. MHD: maternal hypertensive disorder; SDI: sociodemographic index.

**Figure S12.** Age-standardized DALYs rate of MHD for 204 countries and territories by SDI in 2021. DALYs: disability-adjusted life years; MHD: maternal hypertensive disorder; SDI: sociodemographic index.

**Figure S13.** The number of MHD burden across different age groups in different regions from 1990 to 2021. (A) Number of incidence; (B) Number of deaths; (C) Number of DALYs. MHD: maternal hypertensive disorder; SDI: sociodemographic index; DALYs: disability-adjusted life years.

**Figure S14.** EAPC of ASIR, ASMR, and ASDR for MHD from 1990 to 2021 by different age groups and regions. (A) EAPC in ASIR. (B) EAPC in ASMR. (B) EAPC in ASDR. EAPC: estimated annual percentage change; ASIR: age-standardized incidence rate; ASMR: age-standardized mortality rate; ASDR: age-standardized DALYs rate; DALYs: disability-adjusted life years; MHD: maternal hypertensive disorder; SDI: sociodemographic index.

**Figure S15.** Local drifts of MHD incidence with the age, period, and cohort effects by SDI quintiles, 1992−2021. (A) Local drifts of MHD incidence (estimates from age-period-cohort models) for 9 age groups (10−14 to 50−54 years), 1992−2021. The dots and shaded areas indicate the annual percentage change of incidence (% per year) and the corresponding 95% CIs. (B) Age effects are shown by the fitted longitudinal age curves of incidence (per 100,000 person-years) adjusted for period deviations. The dots and shaded areas denote incidence rates or rate ratios and their corresponding 95% CIs. (C) Period effects are shown by the relative risk of incidence (incidence rate ratio) and computed as the ratio of age-specific rates from 1992 to 1996 (the referent period) to 2017−2021. The dots and shaded areas denote incidence rates or rate ratios and their corresponding 95% CIs. (D) Cohort effects are shown by the relative risk of incidence and computed as the ratio of age-specific rates from the 1942 cohort to the 2007 cohort, with the referent cohort set at 1972. The dots and shaded areas denote incidence rates or rate ratios and their corresponding 95% CIs. MHD: maternal hypertensive disorder; SDI: sociodemographic index.

**Figure S16.** Local drifts of MHD deaths with the age, period, and cohort effects by SDI quintiles, 1992−2021. (A) Local drifts of MHD deaths (estimates from age-period-cohort models) for 9 age groups (10−14 to 50−54 years), 1992−2021. The dots and shaded areas deaths the annual percentage change of mortality (% per year) and the corresponding 95% CIs. (B) Age effects are shown by the fitted longitudinal age curves of deaths (per 100,000 person-years) adjusted for period deviations. The dots and shaded areas denote deaths rates or rate ratios and their corresponding 95% CIs. (C) Period effects are shown by the relative risk of deaths (deaths rate ratio) and computed as the ratio of age-specific rates from 1992 to 1996 (the referent period) to 2017−2021. The dots and shaded areas denote deaths rates or rate ratios and their corresponding 95% CIs. (D) Cohort effects are shown by the relative risk of deaths and computed as the ratio of age-specific rates from the 1942 cohort to the 2007 cohort, with the referent cohort set at 1972. The dots and shaded areas denote deaths rates or rate ratios and their corresponding 95% CIs. MHD: maternal hypertensive disorder; SDI: sociodemographic index.

**Figure S17.** Local drifts of MHD DALYs with the age, period, and cohort effects by SDI quintiles, 1992−2021. (A) Local drifts of MHD DALYs (estimates from age-period-cohort models) for 9 age groups (10−14 to 50−54 years), 1992−2021. The dots and shaded areas indicate the annual percentage change of DALYs (% per year) and the corresponding 95% CIs. (B) Age effects are shown by the fitted longitudinal age curves of DALYs (per 100,000 person-years) adjusted for period deviations. The dots and shaded areas denote DALYs rates or rate ratios and their corresponding 95% CIs. (C) Period effects are shown by the relative risk of DALYs (DALYs rate ratio) and computed as the ratio of age-specific rates from 1992 to 1996 (the referent period) to 2017−2021. The dots and shaded areas denote DALYs rates or rate ratios and their corresponding 95% CIs. (D) Cohort effects are shown by the relative risk of DALYs and computed as the ratio of age-specific rates from the 1942 cohort to the 2007 cohort, with the referent cohort set at 1972. The dots and shaded areas denote DALYs rates or rate ratios and their corresponding 95% CIs. MHD: maternal hypertensive disorder; DALYs: disability-adjusted life years; SDI: sociodemographic index.

**Figure S18.** Decomposition analysis of incidence, deaths, and DALYs due to MHD across different regions from 1990–2021. (A) Changes in incidence. (B) Changes in deaths. (B) Changes in DALYs. DALYs: disability-adjusted life years; MHD: maternal hypertensive disorder; SDI: sociodemographic index.

**Figure S19.** Frontier analysis based on SDI and age-standardized incidence rate of MHD from 1990 to 2021 (A) and their correlation in 2021 (B). Black solid lines delineate the frontier line, and black dots indicate the countries and territories. Black labels represent the top 15 countries with the largest effective difference (largest age-standardized incidence rate gap from the frontier). Blue labels represent frontier countries with low SDI and small effective difference (e.g., Somalia, Nepal, Bangladesh, Bhutan, Haiti), while red labels indicate countries and territories with high SDI and relatively large effective difference for their level of development (e.g., United States of America, Lithuania, Germany, Norway, Belgium). A Red dot indicates an increase in age-standardized incidence rate between 1990 and 2021, and a blue dot indicates a decrease. SDI: sociodemographic index; MHD: maternal hypertensive disorder.

**Figure S20.** Frontier analysis based on SDI and age-standardized deaths rate of MHD from 1990 to 2021 (A) and their correlation in 2021 (B). Black solid lines delineate the frontier line, and black dots indicate the countries and territories. Black labels represent the top 15 countries with the largest effective difference (largest age-standardized deaths rate gap from the frontier). Blue labels represent frontier countries with low SDI and small effective difference (e.g., Somalia, Nepal, Lao People’s Democratic Republic, Mali, Bhutan), while red labels indicate countries and territories with high SDI and relatively large effective difference for their level of development (e.g., United States of America, Monaco, Canada, Netherlands, Sweden). A Red dot indicates an increase in age-standardized deaths rate between 1990 and 2021, and a blue dot indicates a decrease. SDI: sociodemographic index; MHD: maternal hypertensive disorder.

**Figure S21.** Frontier analysis based on SDI and age-standardized DALYs rate of MHD from 1990 to 2021 (A) and their correlation in 2021 (B). Black solid lines delineate the frontier line, and black dots indicate the countries and territories. Black labels represent the top 15 countries with the largest effective difference (largest age-standardized DALYs rate gap from the frontier). Blue labels represent frontier countries with low SDI and small effective difference (e.g., Somalia, Nepal, Lao People’s Democratic Republic, Vanuatu, Bhutan), while red labels indicate countries and territories with high SDI and relatively large effective difference for their level of development (e.g., United States of America, Lithuania, Germany, Norway, Belgium). A Red dot indicates an increase in age-standardized DALYs rate between 1990 and 2021, and a blue dot indicates a decrease. SDI: sociodemographic index; DALYs: disability-adjusted life years; MHD: maternal hypertensive disorder.

## What Is New?

We used the latest Global Burden of Disease (GBD) 2021 study to systematically evaluate the trends in maternal hypertensive disorder (MHD) incidence, mortality, and disability-adjusted life years (DALYs) from 1990 to 2021, stratified by age and the sociodemographic index (SDI).

## What Is Relevant?

From 1990 to 2021, the burden of MHD showed a significant decline globally. The disease burden of MHD is negatively correlated with SDI level, with the age-standardized incidence (ASIR), mortality (ASMR), and DALYs rates (ASDR) in low SDI and low-middle SDI regions and countries higher than the global average. In high SDI regions and countries, however, the ASIR and ASDR for women aged 35-54 years exhibited an upward trend, which may be closely linked to prevalent lifestyle risk factors in high SDI countries.

## Clinical/Pathophysiological Implications?

The global burden of MHD has decreased between 1990 and 2021, significant disparities persist, particularly in low SDI regions, requiring targeted interventions such as strengthening healthcare infrastructure and international cooperation to address the burden. In high SDI regions, managing lifestyle risk factors is also crucial in pregnant women with advanced age.

